# Manual curation of TCGA treatment data and identification of potential markers of therapy response

**DOI:** 10.1101/2021.04.30.21251941

**Authors:** Enrico Moiso

## Abstract

The Cancer Genome Atlas (TCGA) represents a valuable source of genomic and clinical data across different tumor types. In this work, TCGA treatment response information have been manually curated in order to be computationally exploited. Drug names were manually corrected to remove typos and to unify commercial and molecule names. Moreover the drug therapy related information has been reorganized in a table to facilitate readability and analysis. The data were benchmarked with uni and multivariable statistical analyses to identify putative markers of response.

## Introduction

The Cancer Genome Atlas (TCGA) project generated more than 2.5 petabytes of genomic and clinical data for 33 different cancer types. While the molecular data were generated by means of bioinformatic pipelines, returning computationally ready to use data, the clinical data were mainly manually annotated, without annotation standards. Therefore, some clinical data are not ready to use from a computational point of view. A clear example is represented by drug names, containing both compound and commercial names, spelled in a variety of, not always correct, ways. The main objective of this study is to report the curation of the treatment-related information of the patients enrolled in the TCGA study, to present the data in an easily accessible format and to show some representative analysis aimed at identifying putative molecular markers of response to various cancer therapies.

## Results

### The manual curation of TCGA treatment response data: the selection of pertinent features and the treatment name harmonization

The TCGA clinical data are organized in dataframes containing clinical features as rows and patients as columns for each of the main 33 tumor type and 5 tumor macro-groups (COADREAD, GBMLGG, KIPAN, STES, LUNG). Refer to the following table for the tumor types present in the TCGA study and their official symbols.

| Acronym | Tumor Type | Acronym | Tumor Type |
| --- | --- | --- | --- |
| ACC | Adrenocortical carcinoma | LUAD | Lung adenocarcinoma |
| BLCA | Bladder Urothelial Carcinoma | LUNG | LUAD + LUSC |
| BRCA | Breast invasive carcinoma | LUSC | Lung squamous cell carcinoma |
| CESC | Cervical squamous cell carcinoma and endocervical adenocarcinoma | MESO | Mesothelioma |
| CHOL | Cholangiocarcinoma | OV | Ovarian serous cystadenocarcinoma |
| COAD | Colon adenocarcinoma | PAAD | Pancreatic adenocarcinoma |
| COADREAD | COAD + READ | PCPG | Pheochromocytoma and Paraganglioma |
| DLBC | Lymphoid Neoplasm Diffuse Large B-cell Lymphoma | PRAD | Prostate adenocarcinoma |
| ESCA | Esophageal carcinoma | READ | Rectum adenocarcinoma |
| GBM | Glioblastoma multiforme | SARC | Sarcoma |
| GBMLGG | GBM + LGG | SKCM | Skin Cutaneous Melanoma |
| HNSC | Head and Neck squamous cell carcinoma | STAD | Stomach adenocarcinoma |
| KICH | Kidney Chromophobe | STES | ESCA + STAD |
| KIPAN | KICH + KIRC + KIRP | TGCT | Testicular Germ Cell Tumors |
| KIRC | Kidney renal clear cell carcinoma | THCA | Thyroid carcinoma |
| KIRP | Kidney renal papillary cell carcinoma | THYM | Thymoma |
| LAML | Acute Myeloid Leukemia | UCEC | Uterine Corpus Endometrial Carcinoma |
| LGG | Brain Lower Grade Glioma | UCS | Uterine Carcinosarcoma |
| LIHC | Liver hepatocellular carcinoma | UVM | Uveal Melanoma |

Fore each tumor type the amount of clinical entries (i.e. sample specific information) available is highly variable, ranging from 634 for UVM samples to 5516 for GBMLGG samples, Figure 1.

**Figure 1:**
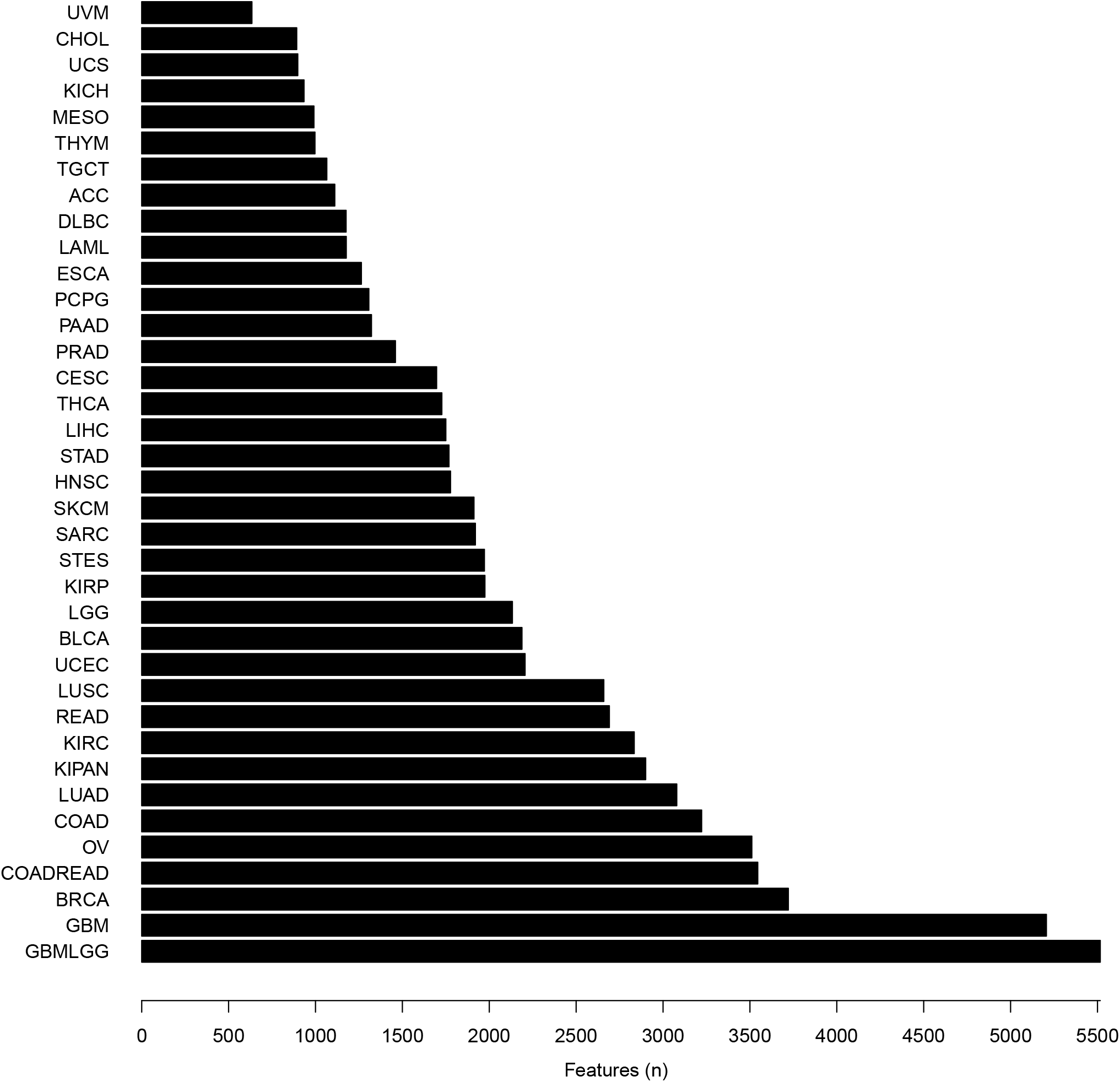
Number of clinical entries per tumor type.

This variable amount of clinical features can be explained by the presence of several features that are cancer specific (i.e. PAM50 classification for BRCA, Gleason score for PAAD) and by the fashion the data are organized. One example is represented by the COAD patient *TCGA-A6-2671* which has been subjected to 22 different treatment, creating 22 different entries for drug related data such as: treatment start, end, response and name. Figure 2 illustrates the list and the hierarchical organization of treatment specific information that has been selected for the manual curation, the radiation data will be covered in different work.

**Figure 2:**
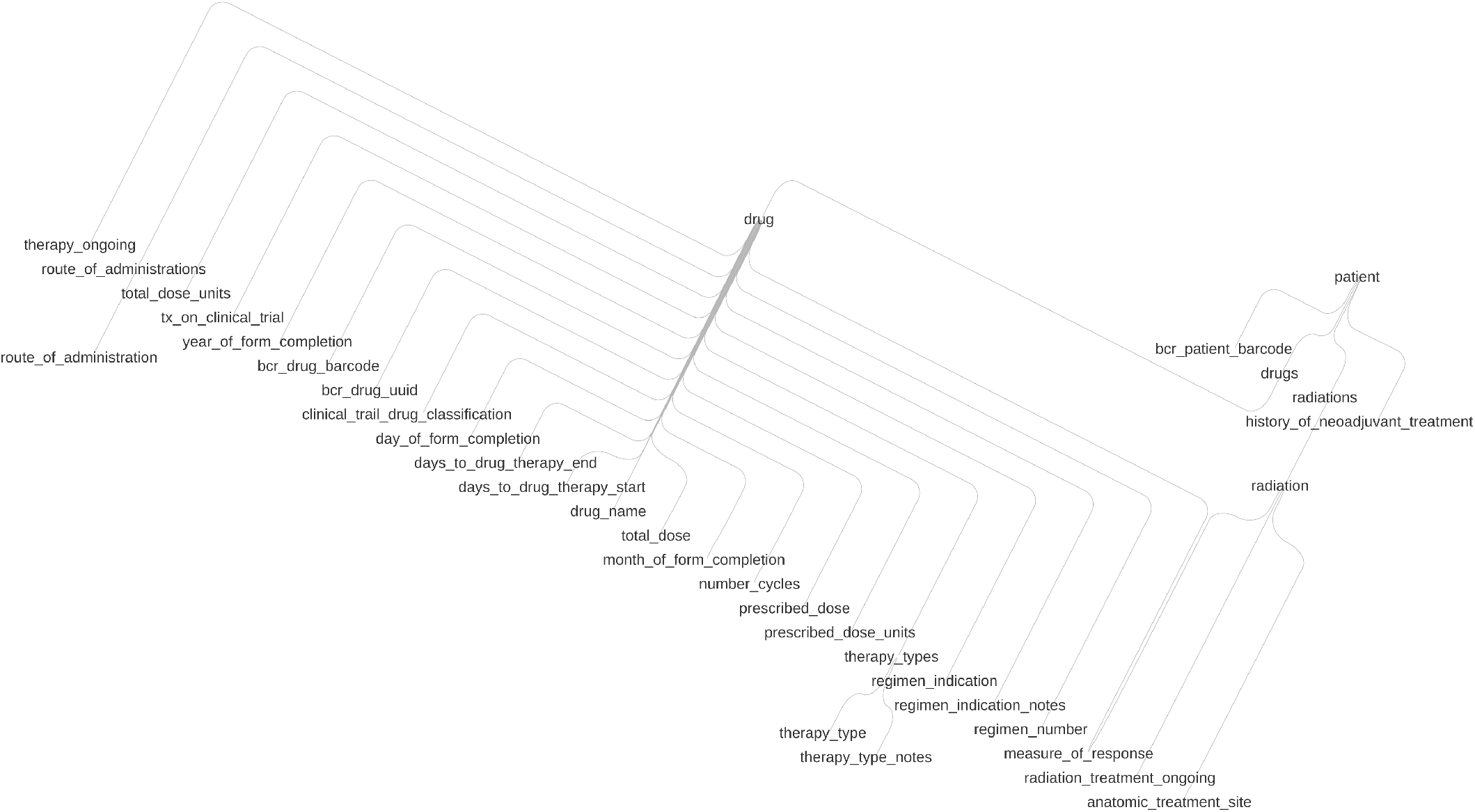
Hierarchical organization of selected clinical features.

The drug name section represents the main obstacle to the systematic analysis of TCGA drug response data. Indeed this information was manually notated, without following any standardization or formal rule. Thus, a drug could have been annotated either with its compound name, commercial name, symbol or abbreviation, moreover several spelling error can be found. For example the drug Pemetrexed, commercial name Alimta^*™*^, has been annotated as Almita and Atima while Fluorouracil was annotated with more than 40 different ways. Figure 3 shows the frequency distribution of all the drug names.

**Figure 3:**
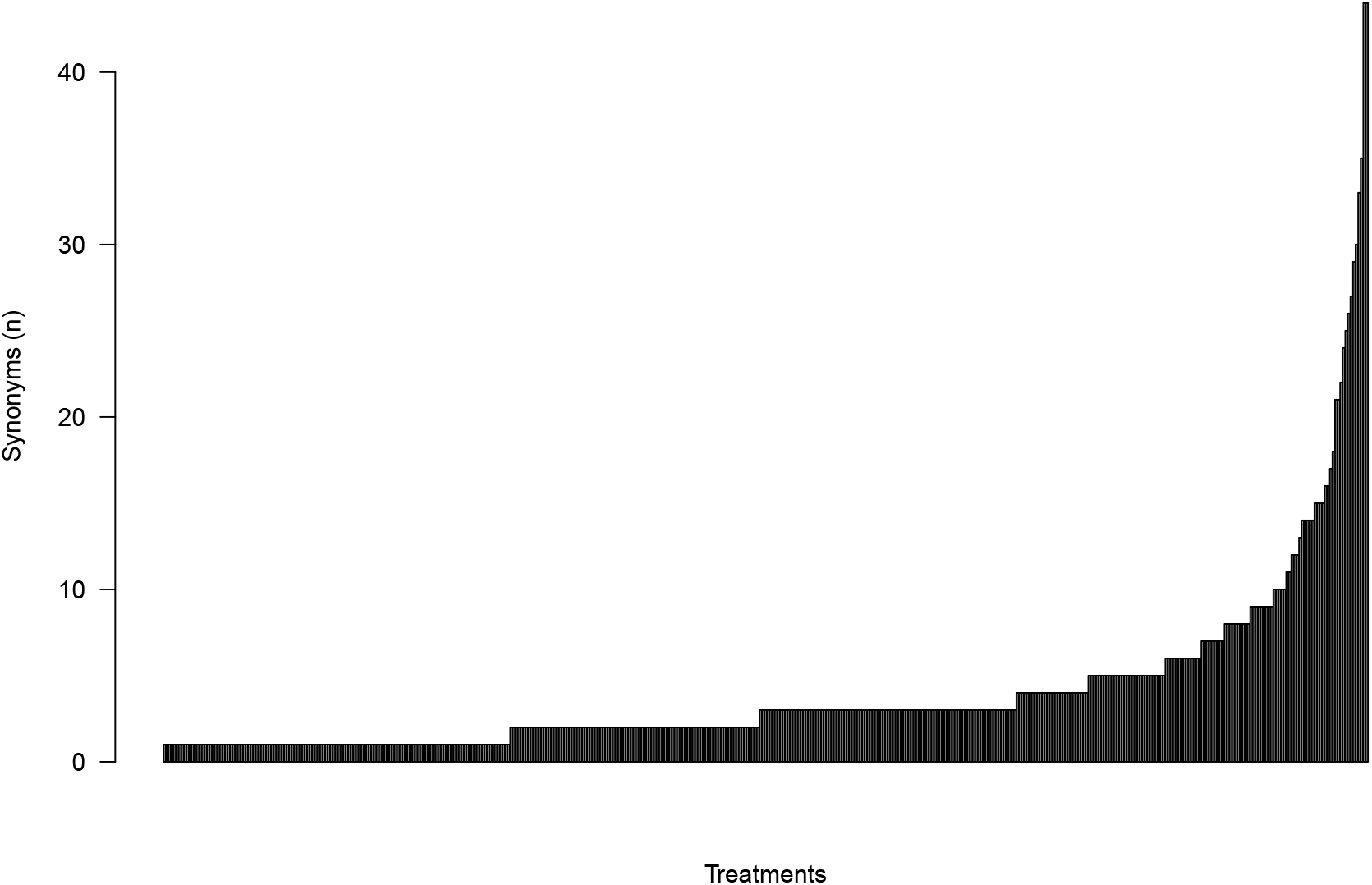
Number of synonyms for every treatment.

As a consequence of the drug name harmonization step, treatment information can be now effectively exploited for statistical analysis. Furthermore the cross-talk with other source of drug related information, such as DrugBank [1] or ClinicalTrials.gov is now possible. Figure 4 and *supplementary data 1*) shows a network representation of drug-target interactions gathered from DrugBank: We can appreciate how drugs cluster on target similarity.

**Figure 4:**
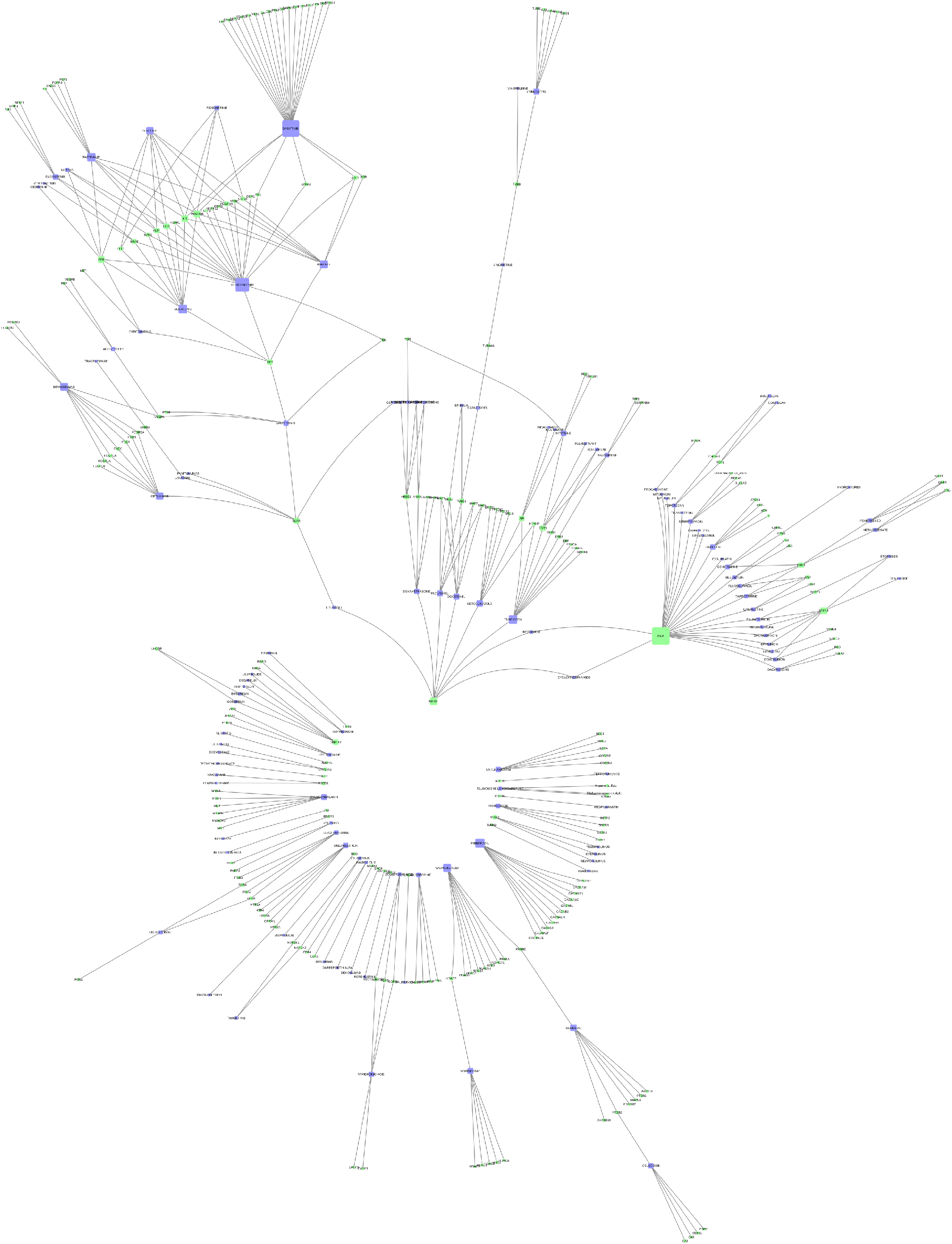
Graph representation of drugs-target interactions for TCGA treatments. Nodes in green represents molecular targets while nodes in purple represents drugs.

The curation steps described so far allowed the creation of the dataframe (drug therapy, *supplementary data 2* and dataframe header description, *supplementary data 3*) that represent the starting point for treatment response analyses that can performed on the TCGA cohort.

### Identification of putative markers of treatment response

In order to identify putative markers of pharmacological treatment response, as proposed in this study, the drug response data required to be further filtered. Only the response to the first, in chronological order, treatment was taken into account; moreover only the most commonly prescribed treatment, for each tumor type, was taken into consideration leaving us with a cohort of 679 unique patients and 15 unique treatments, Figure 5. Finally, the measure of response were collapsed into two categories: Partial and complete response were considered as responders while stable disease and clinical progressive disease were considered non responders. In order to reproduce the following analysis, data in *supplementary data 4* should be used.

**Figure 5:**
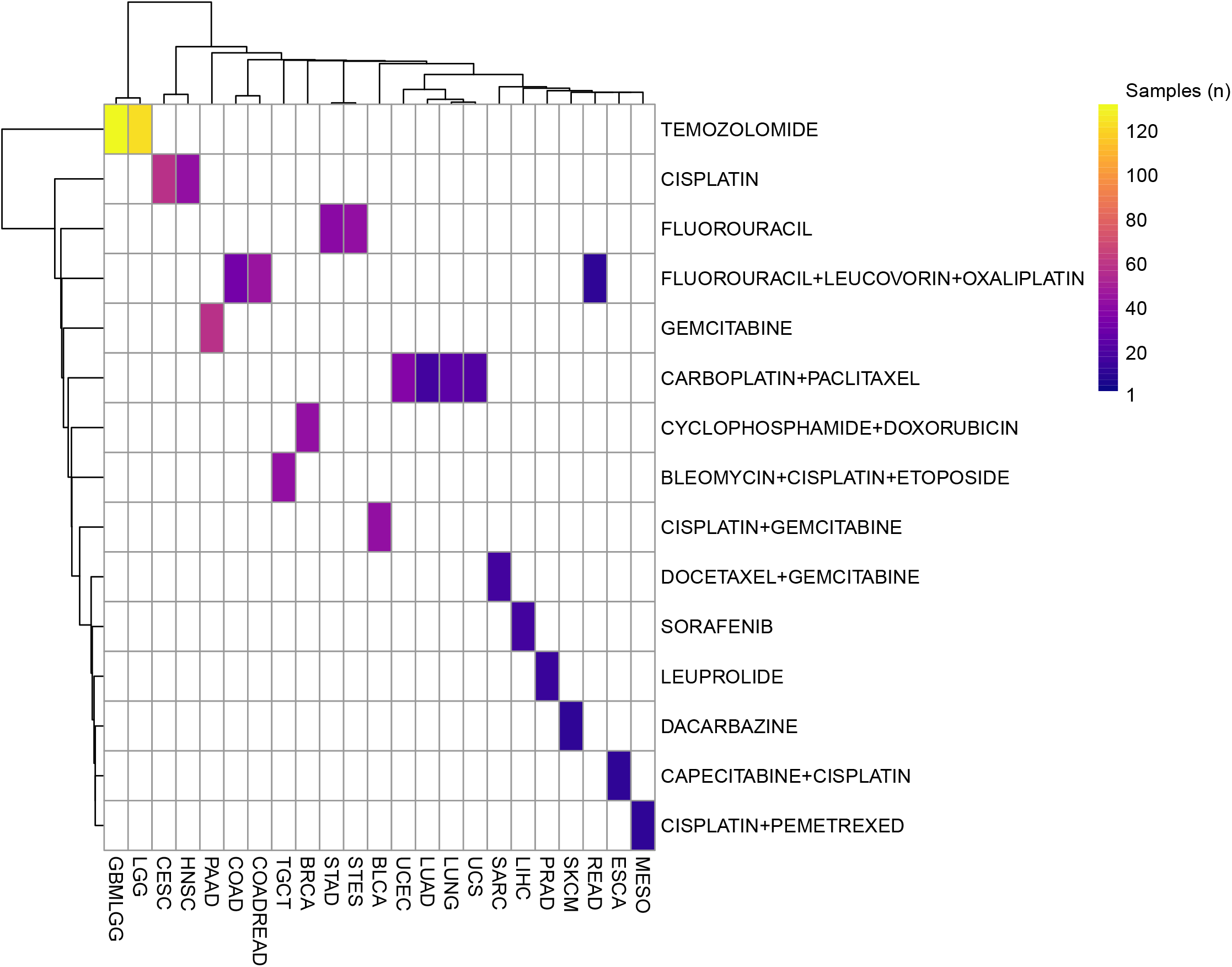
Heatmap showing the number of samples for every treatment (rows) across the various tumor types (columns).

### Correlation of individual features with drug response

The previously generated treatment response data were, at first, systematically correlated with coding gene expression, miRNA expression, protein abundance and clinical data, independently. Figure 6 shows a set of examples of individual feature correlations between three molecular features (miR-10b expression, AGAP21 expression and mTOR phosphorylation in Serine 2448), one clinical feature, (namely histological subtype) and treatment response in 4 different tumor types: CESC, SARC, STES and BRCA, respectively.

**Figure 6:**
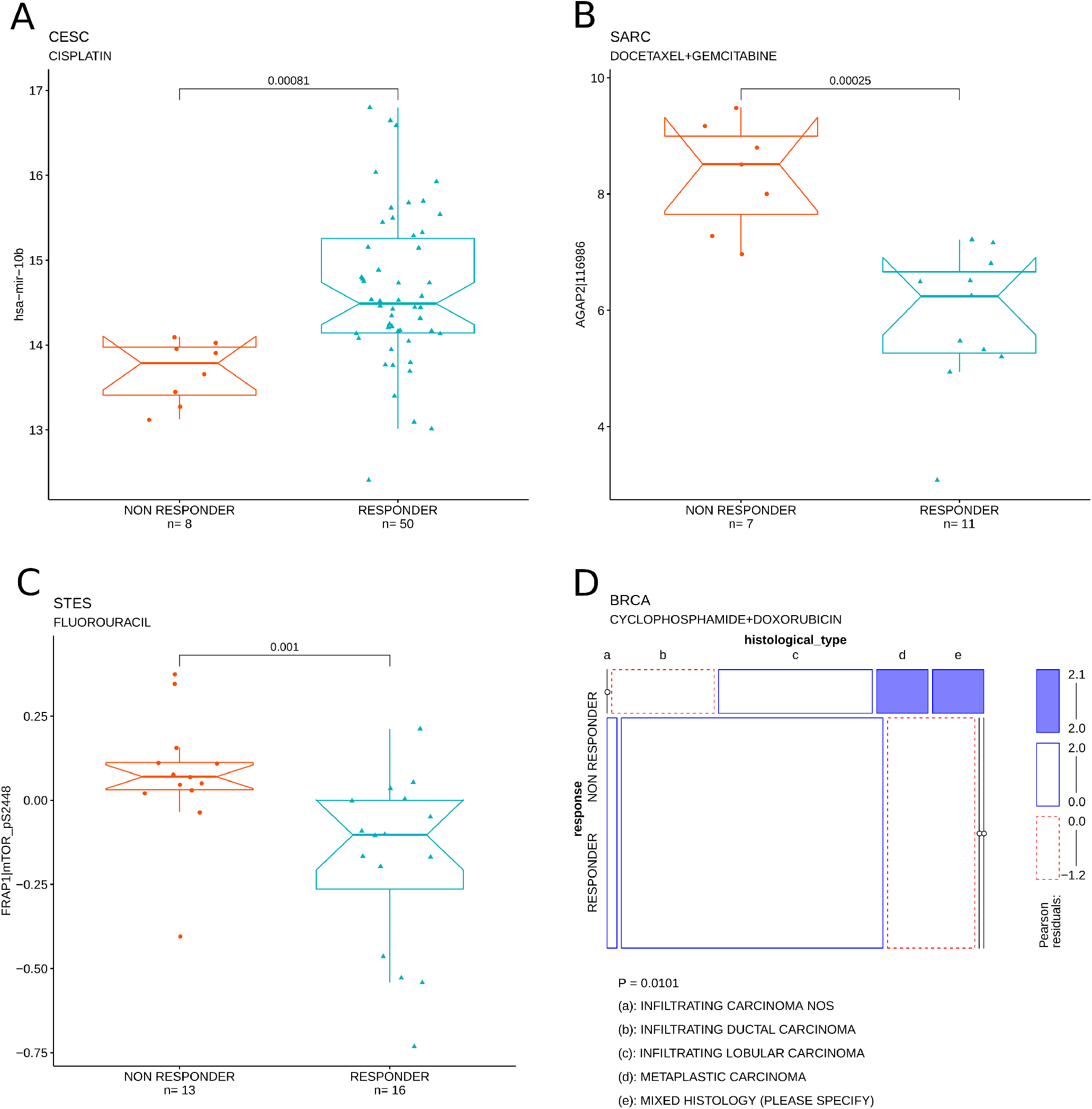
Examples of individual feature correlations with treatment response. A) Correlation between mir-10b, miRNA expression and cisplatin response in CESC. B) Correlation between AGAP1, mRNA expression and response to docetaxal + gemcitabine treatment in SARC. C) Correlation between mTOR phosphorylation, RPPA signal and fluorouracil response in STES. E) Correlation between histological type, clinical feature and cyclophosphamide + doxorubicin treatment in BRCA.

For every tumor type, individual molecular or clinical features have been correlated with the response to the most frequently administered treatment. Figure 7 shows an heatmap of the nominal P-values obtained from the systematic correlation between the individual features and the tumor specific treatment response data. Not surprisingly the features tend to organize in clusters that are tumor/treatment specific. The list of all the correlations thresholded at nominal P-value < 0.05 can be found in *supplementary data 5*. This analysis allows the identification of individual, putative predictors of response to specific therapies.

**Figure 7:**
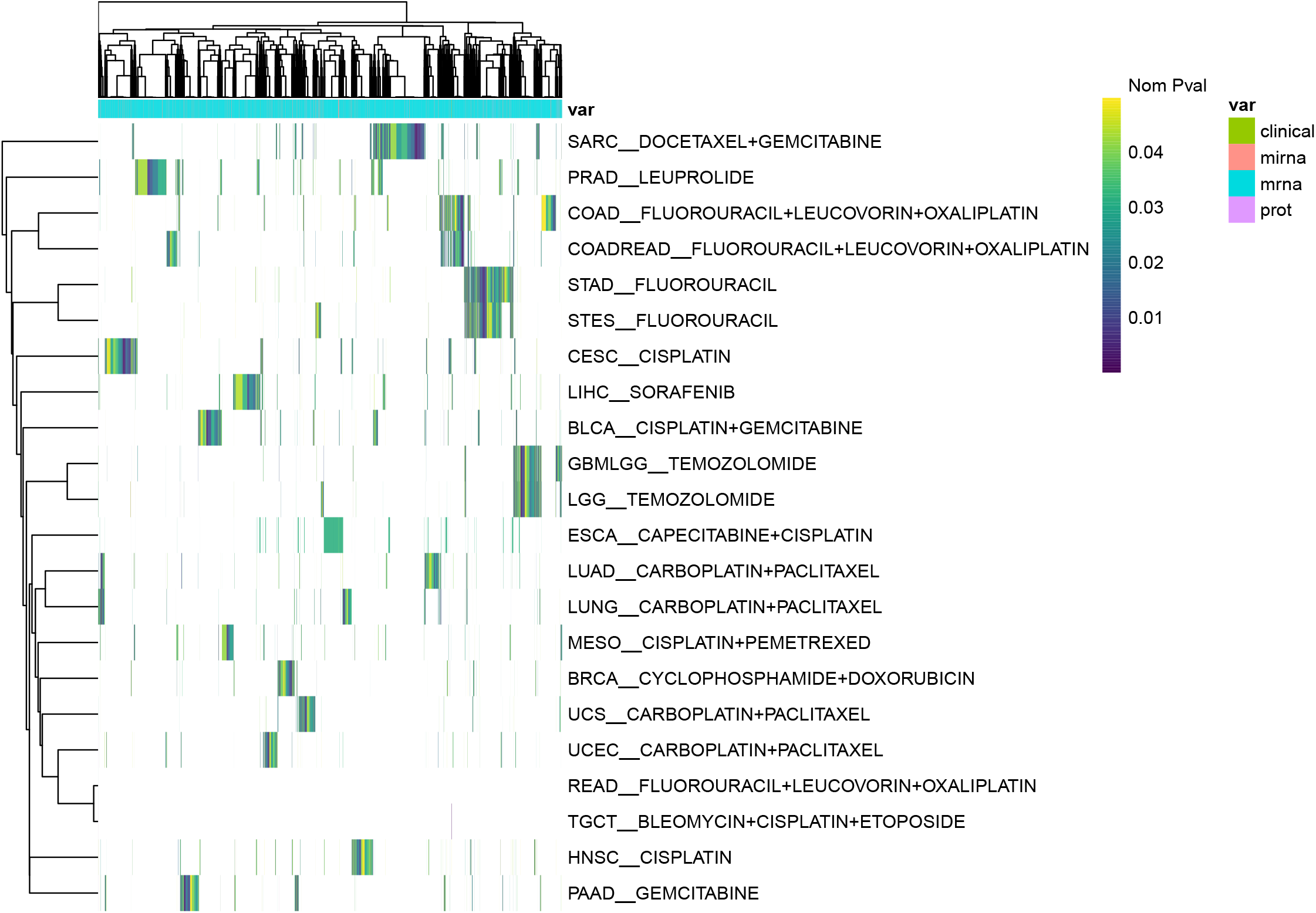
Heatmap summarizing the pvalues of the correlations between molecular or clinical features and the response to the most frequently administered, tumor specific treatment.

Due to the tendency of the molecular features to create response clusters, probably as direct consequence of the intrinsic correlation between genes with similar function [2] or belonging to similar biological pathways, a principal component analysis [3] (PCA) was performed to reduce the amount of the individual molecular features to test. PCA was performed for every tumor types on the set aggregated, most variable features between responders and non responders

The resulting principal components (PCs) were then tested for correlation with drug response data and the results, thresholded at nominal P-value <0.05 are shown in Figure 8. In order to extrapolate the biological meaning of the PCs a Gene Set Enrichment Analysis (GSEA) [6] can be performed on the PCA loadings. In Figure 9 an example of follow up analysis on a PC that correlate with fluorouracil response in STES is shown.

**Figure 8:**
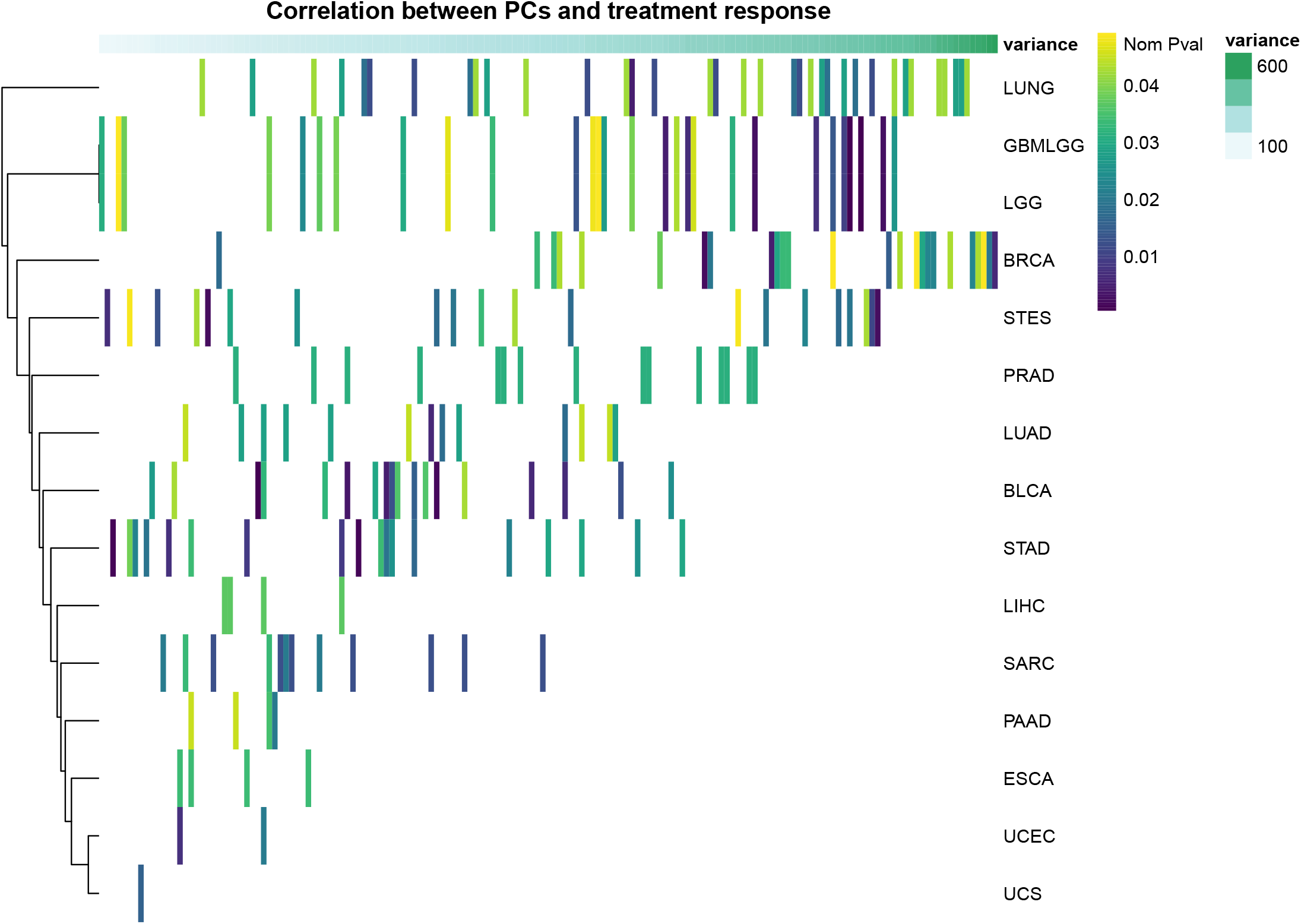
Heatmap summarizing the pvalues of the correlations between principal components (PCs) and the response to the most frequently administered, tumor specific treatment. The components are ordered based the amount of variablity explained.

**Figure 9:**
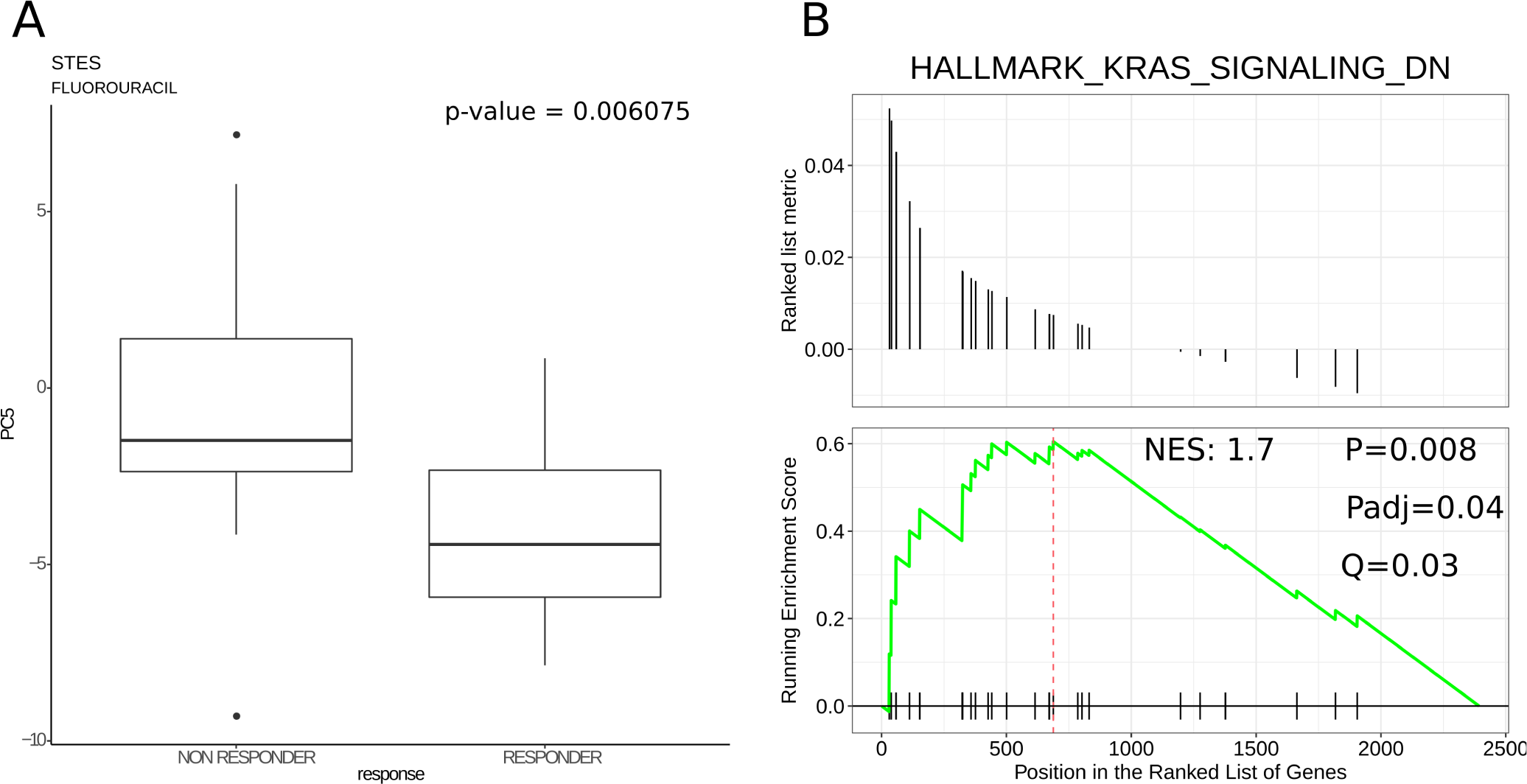
Example of interpretative analysis of a PC that correlates with treatment response. A) PC5 correlates with fluouracil response in STES. B) GSEA analysis on PC5 loading results.

These results show that with these data we can identify individual molecular features and biological processes that correlate with treatment response.

### Multivariate logistic models to predict treatment response

Due to the binary nature or the collapsed response variable, logistic models [8] has been chosen as statistical tool to perform multivariate predictive analysis of response. The logistic models have been build on the combinations of the most strongly response-correlated: 1) molecular features and 2) PCs. In both cases each tumor cohort was split into a training and a test set, the selection of both 1) and 2) as well as the creation of the model was performed on the training set and then validated on the test set. Figure 10 shows the performance of the logistic models on the combined molecular features and PCs on the same graph for direct comparison. The models built on the PCs seem to perform better in the majority of the tested tumor types.

**Figure 10:**
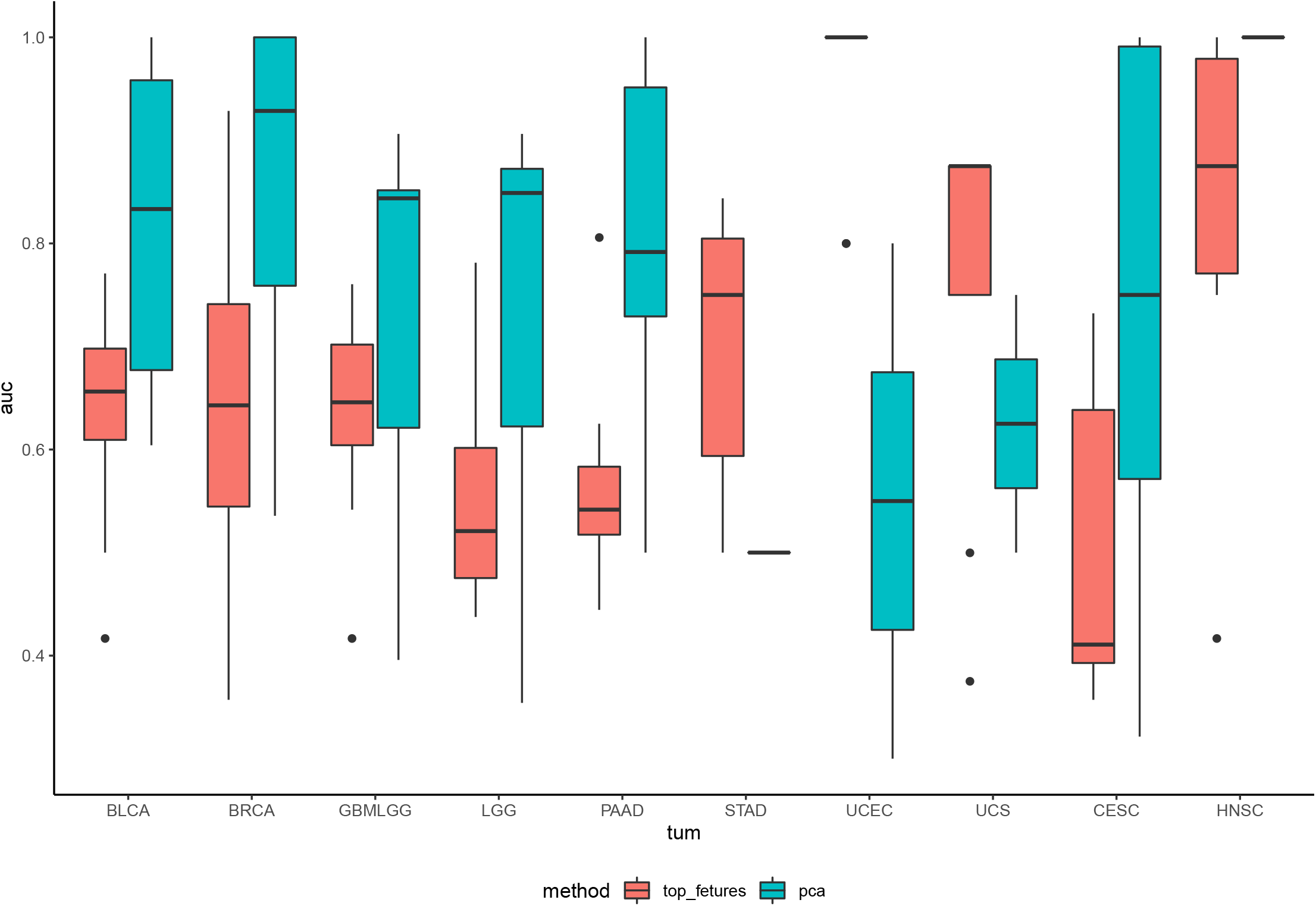
Boxplot representing the accuracy of the logit models built on combined molecular features and PCs. The accuracy is measured by means of the area under the curve (auc) on the testing set.

We also tested the treatment response data by means of a logistic penalized regression, ElasticNet [9]. This represent the hardest test for the dataset since ElasticNet requires cross-validation for tuning method hence a bigger cohort. Therefore we applied ElasticNet only on the GBMLGG cohort which represent the most abundant one. In Figure 11 the results of ElasticNet are shown.

**Figure 11:**
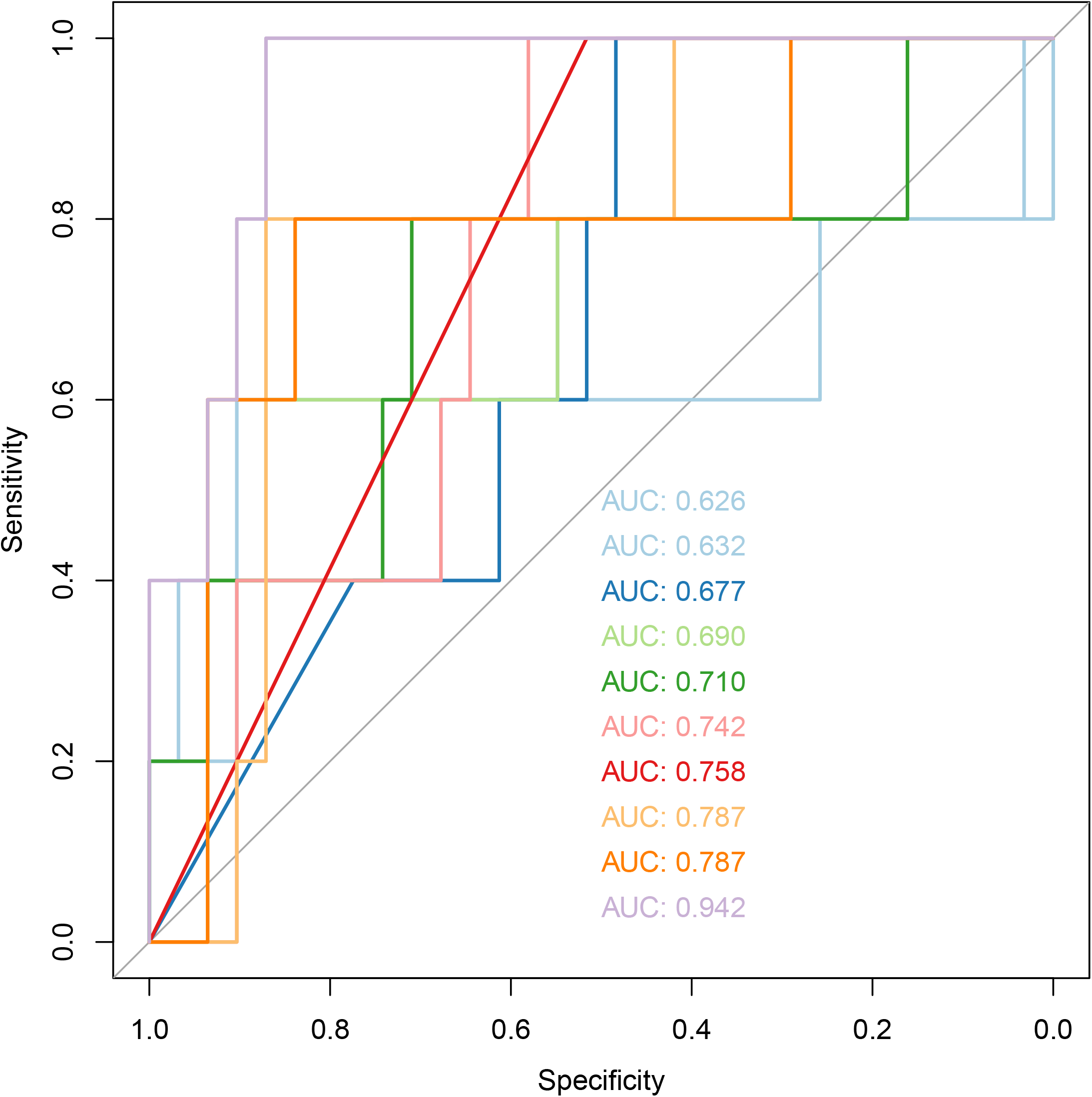
Bidimensional representation of the performance curve of the penalized regression model.

## Discussion

The TCGA represent a formidable resource of molecular and clinical data that allowed an unprecedented comprehension of cancer biology. However the use of TCGA data to study mechanisms and markers of response to therapies has been limited due to the form of the drug response data. In this study we demonstrate that after manual curation the data can be exploited at different levels to identify putative markers of response by means of different statistical approaches. Despite the effort in aggregate the response information (aggregation that can be further achieved by grouping treatments by targets), some tumor types show very few data points and this translate into P-values that won’t be able to survive the multiple test correction. Nevertheless this represent a potent validation tool for drug response studies due to great amount of tumors analyzed and the unprecedented level of molecular characterization, and the reported analysis represent a fraction of the potential of this dataset.

## Materials and Methods

### Manual curation of treatment response data

The following steps were performed in order to generate the data frames containing the details of the response data to radiotherapy and drug therapy for the TCGA samples.

### Data gathering

The following data of TCGA cancer samples and patients (release 2016_02_28) were downloaded from the Broad TCGA GDAC site (https://confluence.broadinstitute.org/display/GDAC/Home), by means of *firehose_get* Version: 0.4.1.

*Clinical data* (Merge_Clinical.Level_1.2016012800.0.0)

*coding genes expression data* (RNAseqV2_RSEM_genes_normalized data_Level_3)

*protein data* (reverse phase protein array, RPPA) RPPA_AnnotateWithGene.Level_3

*miRNAs expression data* (Mirnaseq_illuminhiseq_mirnaseq_bcgsc_ca Level_3_miR_gene_expression) Data were collected for the 33 main tumor types and 5 tumor classes:

- COADREAD: COAD + READ
- GBMLGG: GBM + LGG
- KIPAN: KICH + KIRC + KIRP
- LUNG: LUAD + LUSC
- STES: STAD + ESCA

### Files selection

Merge_Clinical.Level_1.2016012800.0.0 files were unzipped and clin.merged.txt files were used to create the treatment response database. LAML samples were removed from the analysis due to absence of treatment related information.

### Data filtering 1

The clinical data of the remaining samples were initially screened for the following keywords:

- drug
- treatment
- response
- omf (other malignant form)
- patient_barcode
- pathologic

### Data filtering 2

The subset of clinical data retrieved from step 3 was further filtered by frequency, only the categories found in all the 33 tumor types and the 5 classes were considered. The selected clinical parameters are shown in *figure 1*.

### Drug and radiation data splitting and data frame creation

The clinical information was organized into two data frames depending on the type of treatment: drug or radiation. This subdivision was necessary to create uniform data frames. Both categories contain information about patient barcode, tumor type of origin and treatment response. The column names of the data frames are preserved from the initial TCGA files (the typo of the field clinical_trail_drug_classification was not corrected to allow intersection with the original data). Two column names have been added: tumor, to facilitate the sub-setting of the data frame by tumor type and treatment_n, since some patients may have been treated with different drugs beside the initial treatment. The rows only containing sample_barcode, treament_n and history_of_neoadjuvant_treatment information were removed from the data frame. Details of the fields contained in both data frames are summarized in *supp data 1*.

### Drug name correction and harmonization

Drug names present in the drug specific data frame were manually curated in order to correct and harmonize treatment names. Among the most relevant amendment to the drug_name field the change from brand name to generic name, and typo correction. In case of multiple generic names for one drug, the name found in Drugbank was selected, to facilitate the cross-talk between the two databases. This process reduced the number of drug treatments from 859 to 337. The drug name mapping is presented in *supp data 6*.

### Graph representation

The graphical representation of graphs has been performed with the *Cytoscape* software [10].

### Measure of response aggregation

In order to increase the numerosity of the samples in each response group, the indicators of response have been pooled as follows: partial and complete responders were considered as RESPONDERS while stable disease and clinical progressive disease were considered as NON RESPONDERS.

### Single molecular feature correlation

Correlation between continuous variables and measure of response was calculated by means of the Mann-Whitney test. Correlation between categorical variables and measure of response was calculated by means of X^2 test.

### Principal Component Analysis

The PCA was performed on the top 3000, centered and scaled, variable features, that also showed to be individually correlated with the response. Variable features have been identified by coefficient of variation *CV*_*i*_ = *σ*_*i*_*/µ*_*i*_. The PCAs have been calculated by means of *prcomp()* R function.

### GSEA

The GSEA has been calculated on the coding gene component of the PCs loadings. GSEA was performed by means of *clusterProfiler* [11] R package.

### Logistic regression

For what concerns the creation of the logit models on the combination of indivdual molecular features, the following steps have been performed:

1. Select the samples, *s*, with mirna, mrna and protein data available
2. Split the cohort in train and test (70%-30%) sets
3. Identify the most highly response-correlated (assesd by pvalue) features on train test and build model on selected features. To avoid overfitting, a number of features *n* has been selected such as *n* < *s*/10
4. Validate the model on the test set
5. Steps 2-4 are repeated 10 times

For what concerns the creation of the logit models on the combination of PCs, the following steps have been performed:

1. Select the samples, *s*_1_, with mirna, mrna and protein data available
2. Split the cohort in train and test (70%-30%) sets
3. Perform PCA on train set
4. Identify the most highly response-correlated (assesd by pvalue) PCs on train test and build model on selected features. To avoid overfitting, a number of PCs *p* has been selected such as *p* < *s*/10
5. Validate the model on the test set
6. Steps 2-5 are repeated 10 times

The logit regressions have been calculated by means of *lm()* R function.

### ElasticNet

The GBMLGG samples with both mRNA, miRNA and RPPA data were selected and the cohort was divided into training and test seta with the following proportions 70%-30%. The training set was used to tune the elastic net alpha and lambda parameters with a 5 fold cross validation approach and the most accurate model was run on 10 different training and test sets (70/30) of the initially selected cohort to measure the model consistency. In details:

1. Select sample cohort with mirna, mrna and protein data
2. Split cohort in train and test (70%-30%)
3. *α* and *λ* optimization by means of a 5-fold cross-valdiation, cv
4. Split cohort in train and test (60%-40%)
5. Train model with parameters identified in 3
6. Validate the model on test set from 4
7. Steps 4-6 are repeated 20 times
8. The best 10 results have been picked for graphical representation

The ElasticNet models have been calculated by means of *caret* [12] and *glmnet* [13] R packages.

### Statisctical analysis, database creation and plots

The databases have been created by means of custom scripts in BASH and R [14], version 3.6.3. Mann-Whitney test has been used to assess the correlations between continuous variables and treatment response. Pearson’s Chi-squared test has been used to correlate categoric variables for tables bigger than 2×2. Mosaic plots were generated by means of *vcd* [16] R package. Boxplots were generated by means of *ggplot2* [17] R package. ElasticNet result has been represented by means of *ROCR* [18] R package. DrugBank database has been downloaded from https://go.drugbank.com/ in xml format and converted into a dataframe by means of custom scripts.

## Data Availability

The results published here are in whole or part based upon data generated by the TCGA Research Network. The author confirms that the data supporting the findings of this study are available within the article or its supplementary materials.

## Competing Interests

The author declares no conflict of interests

## Grant Information

E.M. was supported by a PhD fellowship of the Italian Ministry of Education, University and Research (MIUR)

## Acknowledgments

The author want to acknowledge Prof. Paolo Provero for critical reading of the manuscript, insightful suggestions and for providing guidance and inspiration. The results published here are in whole or part based upon data generated by the TCGA Research Network: https://www.cancer.gov/tcga.

## Supplementary Material

- Supp data 1: Drug-target mapping
- Supp data 2: TCGA maually curated drug response data
- Supp data 3: database header information
- Supp data 4: filtered, TCGA maually curated drug response data
- Supp data 5: Pvalues of the correlation between individual molecular or clinical features and drug response data
- Supp data 6: mapping between original name and manually curated ones

## Notes

### Competing Interest Statement

The authors have declared no competing interest.

### Summary of Updates

Grammatical text revision, figure 9 revised

